# Eye care risk and safety issues identified by Optometrists in Scotland: a focus group study

**DOI:** 10.1101/2025.07.29.25332365

**Authors:** Dorothy Armstrong, James Graham, Lesley Rousselet, Paul Bowie

## Abstract

**Introduction:** Evidence on the nature and scale of risk and safety concerns in Optometry practice is very limited, especially compared with other primary care professions. In Scotland, many changes occurred in the profession during and because of the Covid-19 pandemic. The theme of risk and patient safety was, therefore, chosen as the priority focus for the 2022 mandatory national training programme, which Optometrists providing General Ophthalmic Services in Scotland are required to undertake.

**Aim:** To explore and identify perceived current and future safety risks and how these might be mitigated in relation to professional Optometry practice in Scotland.

**Methods:** Study participants were all registered General Optical Council (GOC) members from nine of the 14 regional health boards in Scotland. Data was collected via three online focus groups over a 2-month period during 2021 with each comprising six to eight participants. Data were transcribed with permission and then subjected to a basic thematic analysis.

**Results:** 16 Optometrists participated in the study. Six principal themes were generated from the data analysis: 1. Current Overview and Context (e.g. perceived increased risk); 2. Competency Risks (e.g. skills and knowledge to manage disease); 3. Conduct Risks (e.g. behaviours of practitioners); 4. Contextual Risks (e.g environmental issues); 5. Future Risks (e.g. technology); and 6. Risk Mitigations (e.g education and training.)

**Conclusion:** The Optometry profession is concerned the level of clinical risk is increasing, mainly related to technology, scope of practice, role development and changes in patient demand. Multiple recommendations are made to minimise risks including education for new roles; increased focus on improving care quality; support to those involved in safety incidents and complaints; taking a systems approach to areas of high risk; and sharing good practices.

**What is already known on this topic:** *summarise the state of scientific knowledge on this subject before you did your study and why this study needed to be done*

- Research into patient safety and exploring clinical and organisational risks in the primary care professions is underdeveloped compared with secondary care settings.
- In professional optometry practice empirical evidence of safety concerns and risks is even more limited.
- Latterly, there has been some focus on these issues from a professional regulatory, policy, research and education perspective, but this is still rudimentary compared to other primary and community-based professions.

**What this study adds:** *summarise what we now know as a result of this study that we did not know before*

- The findings generated provide some evidence of the reported safety and risk concerns perceived by experienced optometry professionals, particularly related to competency, professional conduct, organisational issues, and future risks. Suggestions for risk mitigations were also captured.

**How this study might affect research, practice or policy:** *summarise the implications of this study*

- The findings provide some insights for policy makers, educators and practitioners on reported safety and risk priorities in the optometry profession.
- Study outcomes have informed the development and implementation of targeted patient safety and risk management education for the optometry profession in Scotland which should be of interest to others in the UK and beyond.
- The findings could also lay the groundwork for a future programme of patient safety improvement and risk reduction interventions similar to those introduced in other primary care settings.

## Introduction

The safety of patient care is a serious public healthcare concern across all healthcare systems and is reportedly one of the leading causes of morbidity and mortality worldwide [1]. Evidence of the prevalence of patient safety incidents (circumstances where patients were harmed or could have been during their interactions with healthcare services) is well-established in acute hospital care – largely because much of the policy attention, and research and resource allocation, has focused in acute due to the perceived greater and more serious risks of harm in this sector. Here, it is estimated that around 10% of patients are unintentionally harmed, with approximately 50% of cases judged to be preventable [2].

In response, a plethora of international and national regulations, policies, educational programmes and improvement initiatives have been developed and implemented over the previous two decades [3–6]. The World Health Organisation (WHO) have initiated a 10-year strategic global action to improve patient safety and also have developed a well-established multi-disciplinary curriculum on the topic [7–8]. The patient safety issue is addressed by regulators as part of the expectations of good clinical practices and is also often included in undergraduate and post-training programmes across different professional groups. In the United Kingdom (UK), for example, we also see the establishment of national patient safety programmes, while it is also integrated within the training curricula of professional bodies and Royal Colleges, often at the behest of clinical regulators [9].

In the primary care professions, research into patient safety occurrences is gradually evolving but is not as extensive or rigorous as the many studies undertaken in secondary care [10]. In general medical practice, a recent systematic review indicated that a safety incident may affect around 2-3% of patients attending face-to-face consultations with clinicians [11]. Comparable safety incident estimates appear to be lacking for community pharmacy and primary care dentistry due the dearth of dedicated research, but each profession has begun to study aspects of the patient safety problem. For example, studies of medication incidents and safety culture in pharmacy are published, as well as professional agreement on some of the most serious types of patient safety incidents that can occur in primary care dentistry [12–14].

In the Optometry profession, the professional body in the UK (College of Optometrists) has published guidance on keeping patients and staff free from harm that covers issues around the quality and safety of care, expected competencies, maintaining trust, and communication and teamwork [15]. However, very little empirical information on the nature of risk and safety concerns is known or published. In a nominal group technique study by researchers in Wales, Optometrists were asked to consider eye-care related safety incidents that they had experience of, or they thought could occur [16]. Incidents related to diagnoses incidents (e.g. related to clinical decision-making) were reported to be infrequent but potentially the most severe, with administration elated incidents perceived the most frequently occurring (e.g. delayed or missed referrals). A group of Australian researchers concluded that rates of burnout amongst Optometrists were higher compared with the general population and other health professionals [17]. In a mixed-methods study by the UK General Optical Council (GOC) to explore risk perceptions and experiences of their registrants, practitioners deemed areas of practice around independent prescribing, detection and management of ocular disease and making appropriate referrals to hold the greatest level of risk [18]. The study highlighted that contextual risks around time constraints, working as a locum and commercial pressures were considered as recurring themes, and ‘most likely’ to occur in practice.

Against this background, the NHS Education for Scotland (NES; Box 1) Optometry team was also aware of the many changes in the profession during and because of the Covid-19 pandemic and therefore the theme of risk and patient safety was chosen as the focus for the 2022 mandatory training programme, which Optometrists providing General Optometry Services in Scotland are required to undertake [19].

#### Box 1. About NHS Education for Scotland (NES) and NES Optometry

NHS Education for Scotland (NES) is an education and training body and a national health board within the National Health Service in Scotland (NHS). It is responsible for developing and delivering healthcare education and training for the NHS, health and social care sector and other public bodies. NES has a Scotland-wide role in undergraduate, postgraduate and continuing professional development.

NES Optometry exists to up-skill community eyecare teams, who provide National Health Service funded care under the General Ophthalmic Services (GOS) terms of service, to enable them to provide high quality eyecare and to improve the ocular health of patients throughout Scotland.

To inform and support the design and focus of the training programme, this preliminary research study was undertaken. The main study aim was to explore and identify the perceived current and future risks and how these might be mitigated in relation to professional Optometry practice in Scotland.

## METHODS

### Definition of Risk

For the purposes of the research, the definition of risk outlined to participants is aligned with the 3-level categorisation out in the GOC report of competency, conduct and contextual risk [20]:

- *Competency risks* – risks from practitioners lacking the necessary skills or knowledge to diagnose and manage disease and conditions or to use appropriate equipment
- *Conduct risks* – risks stemming from the behaviour of practitioners either through negligence or inappropriate behaviour
- *Contextual risks* – features of the environment in which the practitioner operates that may increase the scope for risk or influence the severity or likelihood of clinical and competency risks

### Rapid Search and Review of Literature

A rapid, comprehensive search and review of empirical and ‘grey’ literature was undertaken to identify any key papers to inform the study context and design. The main electronic databases searched over a 15-year period from 1 January 2008 to 31^st^ December 2022 were: PubMed, Medline, SCOPUS, PsycINFO, CINHAL, and Google Scholar. The GOC report and a small sample of published articles of relevance related to risk and patient safety were uncovered. The literature review identified a gap in specific research relating to optometry and risk. Compared to other healthcare professions the data is very much limited to published work by GOC. The 2019 GOC Risk in the Optical professions report (18) found in comparison to other healthcare professions, the roles of Optometrists involve lower levels of risk to patients and the public. However, anecdotally a consensus among practitioners that the level of risk in optical professions is changing required further exploration.

### Study Participants and Setting

Study participants, who were all registered members of the GOC, were voluntarily recruited by invitational email using the NES Optometry organisational database of practising Optometrists providing independent contractor services to the National Health Service (NHS) in Scotland across 14 regional (territorial) health boards.

### Data Collection

The primary method for obtaining data was online focus groups - a qualitative research method whereby a trained facilitator conducts a collective interview of typically between six to ten participants from similar backgrounds and/or similar demographic characteristics of relevance to the study [21]. They were deemed the most appropriate method of data collection as they are useful for exploring perceptions, attitudes and experiences, while enabling participants to debate and challenge each other and clarify their views. The focus groups were facilitated by DA an experienced educator, researcher and healthcare professional and co-hosted by JG, the NES Optometry specialist lead.

Three focus groups were conducted over two months during the spring of 2021. The on-line platform Zoom was used to facilitate the group interviews, and each session was recorded with consent, transcribed and analysed. Focus groups consisted of six to eight people in attendance. Each session lasted around 90 minutes, and we were satisfied that all delegates had the opportunity to participate. Views were also collated using Jam board [an electronic flipchart] and recorded.

### Data Analysis, Interpretation and Validation

Data were subject to a thematic analysis [22] and were deductively categorised using the *a priori* 3-level risk categorisation framework with the addition of themed data related to risk mitigation strategies. The initial coding and generation of categories and themes was undertaken by DA, and these were checked and re-checked by one other researcher (PB) with any disagreements on theme generation resolved through discussion. The two researchers collectively agreed on the selection of quotations that best support the findings related to each of the themes.

## RESULTS

A total of 16 Optometry participants attended three online focus groups. A breakdown of professional and demographic data is outlined in Table 1.

**Table 1.**
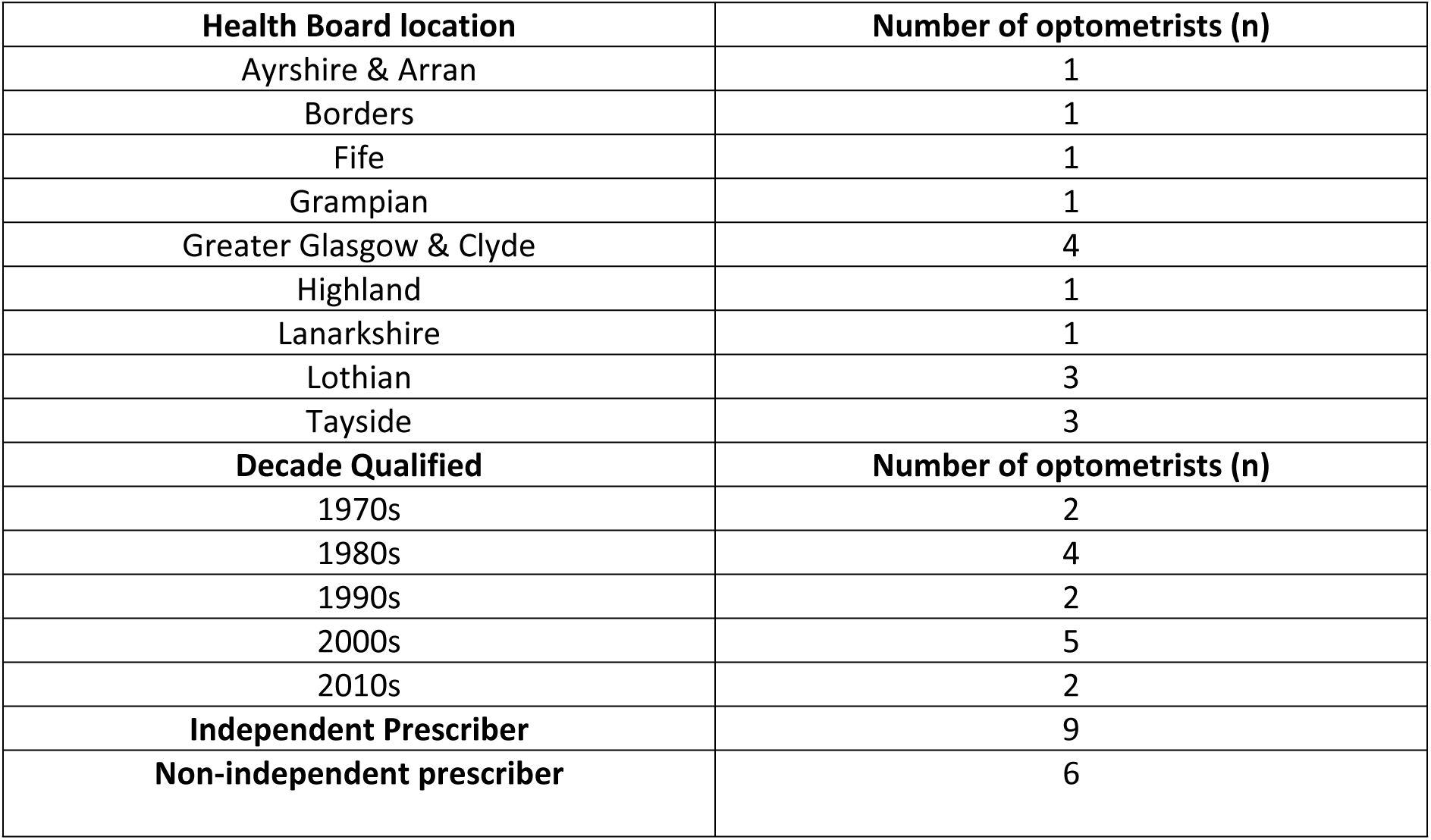
Breakdown of professional and demographic data of study participants (n=16)

| **Health Board location** | **Number of optometrists (n)** |
| --- | --- |
| Ayrshire & Arran | 1 |
| Borders | 1 |
| Fife | 1 |
| Grampian | 1 |
| Greater Glasgow & Clyde | 4 |
| Highland | 1 |
| Lanarkshire | 1 |
| Lothian | 3 |
| Tayside | 3 |
| **Decade Qualified** | **Number of optometrists (n)** |
| 1970s | 2 |
| 1980s | 4 |
| 1990s | 2 |
| 2000s | 5 |
| 2010s | 2 |
| **Independent Prescriber** | 9 |
| **Non-independent prescriber** | 6 |

Six principal themes were generated from the focus group data analysis.

### 1. Current Overview and Context

The perceived levels of current risks varied across the participants. Many highlighted the need for more awareness of known risk factors and education to identify and mitigate the risks. While many agreed with the previous consensus from GOC about the clinical and organisational risks being lower than in other healthcare professions, the majority were clear that these risk areas had increased significantly in previous years. They cited the greater use of complex technology, and expanded roles in prescribing and treatment which, in turn, escalated the risks to ensuring safer patient care. Additionally, the COVID-19 pandemic provided both opportunities and new threats, including the need for a more consistent and robust approach to new ways of working involving the use of technology.

The “First Port of Call” responsibility represents a reframing of the community Optometrist’s remit, intended to promote the understanding that patients’ with ocular complaints should present to an Optometrist in the first instance. The main purpose is to reduce the burden on other primary and secondary care colleagues, since many of these complaints can be safely managed by community optometry practices. First port of call was a common theme which provoked much comment and debate. While the initiative was generally welcomed and seen as a driver for improvement in patient care and treatment, multiple barriers and challenges were identified and discussed, which could be categorised in all three areas of risk i.e. competency linked to scope of practice, conduct linked to communications and behaviours and context linked to the environment, technology and systems.

The logistics around providing 24-hour optometry cover and communication to patients and indeed between healthcare professionals may cause confusion and uncertainty. Some Optometrists were concerned about duplication or overlap with other acute or emergency services such as minor injuries and emergency departments.

“*First Port of Call, …difficult to manage particularly in a small practice…*” Focus Group 1 Participant

### 2. Competency Risks

Several participants noted the high-profile legal case of Honey Rose where a qualified Optometrist was convicted of gross negligence misconduct [23]. Although the conviction was later overturned on appeal, the incident remained a source of concern and anxiety for participants. A common competency risk identified was related to missed diagnoses and a failure to detect disease. Although this was generally thought to be a low risk and unlikely to occur, it was felt that if there was a failing then the consequences would be significant for patients and the practitioners.

A further emerging theme was the risk to patients related to Optometrists who were qualified as independent prescribers of medicines. There was clear concern and worry from participants about the increased pressure in prescribing and the need to ensure full and accurate record keeping and that clinical decision making was recorded in a robust and transparent manner. However, there was agreement that time pressures and inadequately designed work systems sometimes made this challenging for practitioners.

When safety and other incidents do occur, participants agreed that the profession should be encouraged to record and report any ‘near misses’ or adverse events. However, many felt they were not well equipped for this aspect of their role and welcomed the mandatory training in 2022 related to risk and patient safety.

All participants welcomed education and training, however several voiced concerns about elements of professional development being unsympathetic to the varying abilities of practitioners, from novice to expert. More specifically, in an enhanced role such as prescribing or invasive treatments, ongoing mentorship, peer support and clinical audit were considered necessary following the theoretical input.

“…*scope of professional practice has to be matched by training and interaction with secondary care*” Focus Group 3 participant

### 3. Conduct Risks

Discussion centred around risks that derived from conduct issues which was suggested as most often resulting from miscommunication or inappropriate behaviours. Incivility or inappropriate professional behaviour was also discussed as a risk by participants. Concerns around the risks associated with delegating tasks to support staff, including visual field testing, fundus photography and contact lens insertion and removal were highlighted. Participants noted interactions between optometrists and patients, as well as between optometrists and colleagues or other healthcare professionals, all carried risk through inadequate communication.

Professional behaviours were highlighted in relation to maintaining a balance between business needs and delivery of high-quality care. Again, the risks here are to patient safety and to the well-being of the individual optometrists and colleagues. Managing a concern or a complaint openly and honestly and awareness of Scotland’s Duty of Candour legislation was a common theme. It was noted that for individual practitioners to respond effectively to complaints, it is important they have the knowledge, skills and confidence to do so, especially in preventing practitioners becoming overly defensive in their future practice. However, multiple mentioned their lack of knowledge about the legislation and wanted to raise awareness through more focused education and training, including on taking a human factors systems approach to learning from safety events and complaints.

Other issues such as lack of information given or unnecessary delays may lead to challenging conversations with patients and occasionally patients may decide to raise a complaint. Participants reported challenges in responding to patient expectations, particularly during the pandemic: for example, patients referred by general practitioners for minor injuries who had high expectation to be seen urgently.

“*We need a culture of being open and honest, sharing mistakes and discussing with peers*” Focus Group 2 Participant

“*Practitioners going beyond the scope of their practice or competence, increased risk of undermining public confidence in the profession*” Focus Group 3 Participant

### 4. Contextual Risks

The environment Optometrists are working was thought to play a key part in the degree of risk perceived. The variation in risk between working in a rural practice compared to a hospital clinic was highlighted. Multiple variables were highlighted which influenced the environment, for example: the current changes to ways of working, amended policies around patient contact and interactions; new technology such as video and phone consultations have all elevated the degree of concerns and may result in increased risks to both patients and to the well-being of practitioners.

Participants discussed the use of technology and access to remote consultation as both an opportunity and a threat. Variations across Scotland was highlighted and could result in “a postcode lottery” of clinical care. Recall and referral systems were discussed by participants who identified this as a potential high-risk area. There was significant variation across Scotland and appeared to be no defined or recognised good practice. Despite many practices using sophisticated patient management systems, often there is no standard procedures for recalling patients if they did not attend an appointment. Similarly, follow up visits or referrals to secondary care were mixed.

Lone or working in isolation, specifically in remote and rural areas was identified as being a risk to individual practitioners: opportunities for a broader range of patients, access to peer review, education and training was considered less available. Crucial to the success of the newer ways of working is education and training. Education and training provided by NES Optometry was considered to be a positive way of sustaining a workforce who were competent from registration through to enhanced and advanced roles.

“*The retail / clinical division is going to get greater and there will be some Optoms caught in the middle*” Focus Group 1 Participant

### 5. Future Risks

Many of the themes identified relate to the rapid changes to care processes and systems due to the Covid-19 pandemic. Participants agreed that Optometrists responded quickly and effectively to this complex and dynamic crisis situation; however, this may well have inadvertently contributed to the perception of increasing clinical risks by the profession. Participants also shared their concerns about time constraints and balancing the many conflicting requirements in their roles. Keeping up to date with technological advances and new equipment was identified as an opportunity for improved quality of care yet was also a source of stress for some practitioners. Changes in demographics and the shift to community care models are all playing their part in how Optometry professionals gauge future working and hence consider future risks to practice.

*“…scope of professional practice has to be matched by training and interaction with secondary care…”* Focus Group 2 Participant

“*The profession becoming split between those wanting to extend clinical remit and those who do not*…” Focus Group 2 Participant

### 6. Risk Mitigations

Discussion centred around how the Optometry profession has undergone significant changes due to new and enhanced ways of working introduced as part of the response to the Covid-19 pandemic. Participants agreed that these factors have increased the current and perceived risk to patient safety. Multiple approaches to mitigating these risks were suggested by participants (Table 2).

**Table 2.** Risk Categorisation and Suggested Mitigation Strategies.

| Competence Risk | Suggested Mitigation |
| --- | --- |
| Missed diagnosis or failure in treatment | Education and training in risk and patient safety<br>On-going requirement to demonstrate skills and expertise to remain registered |
| Independent prescribing | Education and training opportunities<br>Peer support and review<br>Clinical audit |
| Awareness of risk factors and mitigation | Education and training in risk and patient safety |
| Skills acquisition | Continuing professional development<br>Peer support and review<br>Clinical audit |
| Conduct Risk | Mitigation |
| Poor or inadequate communication with patients and between professionals | Education and training in communication skills<br>Increased awareness of complaints resolution and Duty of Candour |
| Professional behaviours | Mentoring and peer support<br>Feedback and appraisal<br>Clinical audit |
| Managing complaints | Peer support and review<br>Education and training opportunities |
| Context Risk | Mitigation |
| New ways of working including video & 'phone consultations | Education and training in new technology and standards for practice<br>On-going requirement to demonstrate skills and expertise to remain registered |
| Lone or working in isolation | Access to peer review<br>Access to education and training opportunities<br>Access to NES mandatory training |
| Variation in standards to recall and refer systems | Consider national standards / operating systems<br>Clinical audit |

It was agreed that education and training across all sectors of the profession will play a key role in acknowledging, recording and responding to risk. Particularly in relation to continuing professional development, and on-going access to high quality educational opportunities from registration to advanced practice roles. GOC and NHS Education for Scotland (NES) were both applauded for their position in supporting individuals and the optometry profession. Specific education about risk management including complaints and adverse events would be welcomed as is the NES approach to mandatory training. A continuing emphasis on learning and development in relation to leadership and professional behaviours was also suggested as a means to reduce conduct risks. Building on good practice and finding effective ways to disseminate new and innovative working should continue and again the NES mandatory training provides an effective way to manage this. Clinical audit, feedback and appraisal mechanisms were recognised as an important method of improving practice. Adopting robust approaches to mentoring and peer review was also considered a useful and productive way forward.

## DISCUSSION

We have introduced the perceptions of risk in optometric practice in Scotland: gathered as part of our focus group discussions, ahead of developing an educational resource around risk and patient safety in optometry. Aside from the ‘Risk in the Optical Professions’ paper, by the General Optical Council (18) and a recent study by MacFarlane and colleagues [16] research in this area is underdeveloped, especially in comparison to other primary care professions. Our findings make a small contribution to understanding in eye care safety in Optometry practice. They also offer important and previously unreported insights from practitioners around suggestions to enhance current risk mitigation strategies and perceived future risks to safety that may need to be considered.

Within Scotland, there are a significant number of independent prescribing optometrists managing a variety of conditions within their community practices. Amongst those practitioners, there is of course variation in scope of practice, as well as confidence in managing certain conditions. Whilst the expanding remit of the community optometrist is beneficial to patients, as well as reducing impact on other primary care professions and secondary care ophthalmology departments, practitioners within the focus groups shared a feeling of increasing risk in their role. It was highlighted that the wider practice team, and the system which practitioners work within, play an important role in the delivery of safe and effective patient care. Participants in the focus groups shared collective views that they, the Optometrist, believed that the ultimate responsibility of any patients entering the practice resided with them, rather than the overall ‘system’ or practice they are working within. Concerns regarding the ability to oversee every patient interaction were raised.

The findings identified multiple concerns with patient safety and clinical risk issues in professional practice. These contribute to the development of targeted education to address the patient safety culture in community optometric practice: by increasing awareness of clinical risk, and how to manage it more effectively. The aim of this education was to support practitioners in reflecting more on risk in their own practice/work setting and introduce new theory and approaches that will help support a new culture around risk management in optometric practice is perceived and undertaken.

The move to ‘systems thinking’ is well described in a paper by Clarkson et al [24] which recognises healthcare is a highly complex and interacting system of people, equipment, processes, and institutions working together. This “systems perspective” of healthcare can reframe our understanding as to how care is understood, delivered and can be improved and help individuals and professions think differently about risk and patient safety. It is clear that integration of this approach would modernise and benefit quality, safety and risk initiatives and related education in the field of optometry,

A Report by the Professional Standards Authority (2019) recognizes there is no easy solution to complex safety issues and rather regulators, professional bodies, providers and education bodies need to work together. Specific training in risk management, including responding to complaints and adverse events, is welcomed as is the NES approach to mandatory training. A continuing emphasis on learning and development in relation to leadership and professional behaviours will help to reduce conduct risks. Building good practice and finding effective ways to disseminate new and innovative working should continue and again the NES mandatory training provides an effective way to manage this. Clinical audit, feedback and appraisal mechanisms were recognised as an important method of improving practice. Adopting robust approaches to mentoring and peer review was also considered a useful and productive way forward.

### Strengths and Limitations

Focus groups create a safe confidential space where individuals share their views in a dynamic structured conversation between participants. The focus groups offer powerful insights into people’s feelings and thoughts and thus a more detailed, nuanced, and richer understanding of their perspectives on the research topics. We believe the experienced facilitators were able to manage the groups to enable these goals to be met. Limitations included the use of convenience sampling and a lack of professional representation from six regional health boards. The use of Microsoft Teams to conduct the focus groups may have inhibited the flow of discussion to a certain extent, however many participants also contributed comments and ideas via the Chat function.

## CONCLUSION

The optometry profession has undergone significant changes due to new and enhanced ways of working introduced as part of the response to the Covid-19 pandemic. The data generated by this study found those factors have increased the current and perceived risk to patient safety. Education and training across all sectors of the profession will play a key role in acknowledging, recording and responding to risk. Particularly in relation to continuing professional development, on-going access to high quality educational opportunities from registration to advanced practice roles, is paramount. GOC and NHS Education for Scotland were viewed positively for their position in supporting individuals and the optometry profession. Overall, further and deeper research is required to better understand the nature and management of risks in optometry practice. Exploring a targeted safety improvement programme, like those in other primary care professions, may also be beneficial.

## Data Availability

All data produced in the present study are available upon reasonable request to the authors

## Acknowledgments

We like to offer sincere thanks to all Optometry colleagues in Scotland who gave freely of their time to attend and contribute to the online focus groups.

## Ethical review

Under United Kingdom research ethics arrangements this study was considered to be service evaluation involving National Health Service staff and therefore does not require ethical review.

## Consent to participate

Informed consent was obtained using electronic mail communication, which participants read before agreeing to participate in the study. This email outlined the study purpose and assured potential participants that taking part was strictly done on a voluntary basis and that they could withdraw at any stage. Anonymity and the confidentiality of responses was guaranteed for those who consented.

## Data availability statement

The datasets used and/or analyzed during the current study are available from the corresponding author upon reasonable request.

## Patient and Public Involvement

The study did not involve patients or the public but was rather exploring safety issues with the clinical workforce

## Appendix 1 Focus Group Information

Questions for Focus Groups

Facilitated by X and hosted by Y.

1. The GOC in their ‘Risk in the optical professions’ report of 2019 states

“In comparison to other healthcare professions, it is widely accepted that the roles of Optometrists involve lower levels of risk to patients and the public.” (GOC, 2019).

What do you think are the key risks to

- Individual optometrists

- The optometry profession

- Public / patients

Please consider current and future practice.

2. How might we mitigate against the key risks? GOC (2019) suggests

“education, training and qualifications, where practitioners are required to complete additional training or gain additional qualifications before they can work in potentially riskier areas of practice.” (GOC 2019). What are your thoughts?

3. The final question will explore your thoughts using a SWOT analysis on Jam board. Please consider all possible strengths, weaknesses, opportunities and threats in relation to risk and optometry in Scotland.

The discussions will be recorded and transcribed. Individual identities will be anonymised. The Jam board post-its are also anonymised.

## REFERENCES

[1] World Health Organisation. Patient safety. Available from: https://www.who.int/news-room/fact-sheets/detail/patient-safety. Accessed 21 December 2024.

[2] Vincent C, Neale G, Woloshynowych M. Adverse events in British hospitals: preliminary retrospective record review. BMJ 2001;322:517–9

[3] The Joint Commission. National Patient Safety Goals. Available from: https://www.jointcommission.org/standards/national-patient-safety-goals/. Accessed 21 December 2024.

[4] Scottish Patient Safety Programme. https://ihub.scot/improvement-programmes/scottish-patient-safety-programme-spsp/. Accessed 21 December 2024.

[5] Canadian Patient Safety Program. https://chalearning.ca/programs-and-courses/canadian-patient-safety-program/. Accessed 21 December 2024.

[6] Australian Commission on Quality and Safety in Health Care. https://www.safetyandquality.gov.au/. Accessed 21 December 2024.

[7] World Health Organisation. Resolution WHA55 18. Quality of care: patient safety. In: Fifty-fifth World Health Assembly, Geneva, 13–18th May 2002. Available from: https://apps.who.int/gb/ebwha/pdf_files/wha55/ewha5518.pdf Accessed 21 July 2024.

[8] World Health Organisation. Global patient safety action plan 2021–2030. Available from: https://www.who.int/teams/integrated-health-services/patient-safety/policy/global-patient-safety-action-plan. Accessed 21 December 2024.

[9] Academy of Medical Royal Colleges. National Patient Safety Syllabus Launched to Save Lives. https://www.aomrc.org.uk/publication/national-patient-safety-syllabus-launched-to-save-lives/. Accessed 21 December 2024.

[10] Verstappen W, Gaal S, Bowie P, Parker D, Lainer M, Valderas JM, Wensing M, Esmail A. A research agenda on patient safety in primary care. Recommendations by the LINNEAUS collaboration on patient safety in primary care. Eur J Gen Pract. 2015 Sep; 21 Suppl(sup1):72–7.

[11] Panesar SS, deSilva D, Carson-Stevens A, Cresswell KM, Salvilla SA, Slight SP, Javad S, Netuveli G, Larizgoitia I, Donaldson LJ, Bates DW, Sheikh A. How safe is primary care? A systematic review. BMJ Qual Saf. 2016 Jul;25(7):544–53.

[12] Black I, Bowie P. Patient safety in dentistry: development of a candidate ’never event’ list for primary care. Br Dent J. 2017 May 26;222(10):782–788.

[13] Ensaldo-Carrasco E, Sheikh A, Cresswell K, Bedi R, Carson-Stevens A, Sheikh A. Patient Safety Incidents in Primary Care Dentistry in England and Wales: A Mixed-Methods Study. J Patient Saf. 2021 Dec 1;17(8):e1383–e1393.

[14] Adie K, Fois RA, McLachlan AJ, Walpola RL, Chen TF. The nature, severity and causes of medication incidents from an Australian community pharmacy incident reporting system: The QUMwatch study. Br J Clin Pharmacol. 2021 Dec;87(12)

[15] College of Optometrists. Protecting patients, colleagues and others from harm. https://www.college-optometrists.org/clinical-guidance/guidance/safety-and-quality/protecting-patients,-colleagues-and-others-from-ha. Accessed 21 December 2024.

[16] MacFarlane E, Carson-Stevens A, North R, Ryan B, Acton J. A mixed-methods characterisation of patient safety incidents by primary eye care practitioners. Ophthalmic Physiol Opt. 2022 Nov;42(6):1304–1315.

[17] Bentley, SA and Black, A and Khawaja, N and Fylan, F and Griffiths, AM and Wood, JM (2021) The mental health and wellbeing survey of Australian optometrists. Ophthalmic and Physiological Optics. ISSN 1475–1313 DOI: 10.1111/opo.12823

[18] General Optical Council. Risk in the optical professions final report. July. 2019. Available from: https://optical.org/media/55zjedes/risk-in-the-optical-professions-2019.pdf Accessed 21 December 2024.

[19] GENERAL OPHTHALMIC SERVICES (GOS) AND NHS OPTICAL VOUCHERS – LEGISLATION CHANGES. https://www.publications.scot.nhs.uk/files/pca2022-o-03.pdf Accessed 21 December 2024.

[20] General Optical Council. Standards for optometrists and dispensing opticians. Available from: https://optical.org/optomanddostandards/ Accessed 21 December 2024.

[21] . Kitzinger J. The methodology of focus groups: the importance of interaction between research participants. Sociology of health & illness. 1994 Jan;16(1):103–21.

[22] Braun V, Clarke V. Thematic analysis. In Encyclopedia of quality of life and well-being research 2024 Feb 11 (pp. 7187-7193). Cham: Springer International Publishing.

[23] British Broadcasting Corporation News. Vincent Barker death: Optometrist Honey Rose conviction quashed. https://www.bbc.co.uk/news/uk-england-suffolk-40776091 Accessed 21 December 2024.

[24] Clarkson J, Dean J, Ward J, Komashie A, Bashford T. A systems approach to healthcare: from thinking to -practice. Future Healthc J. 2018 Oct;5(3):151–155. doi: 10.7861/ futurehosp.5-3-151. PMID: 31098557; PMCID: PMC6502599.

